# Peripheral Blood DNA Methylation Changes after Omega-3 Fatty Acid Treatment Indicate Anti-inflammatory Effects and Individual Variability

**DOI:** 10.1101/2020.10.09.20209726

**Authors:** David E Frankhouser, Sarah Steck, Michael G Sovic, Martha A Belury, Ralf Bundschuh, Pearlly S Yan, Lisa D Yee

## Abstract

**Background:** Omega-3 or n-3 polyunsaturated fatty acids (PUFAs) are widely studied for health benefits based on potential anti-inflammatory effects. However, the factors involved in mediating the anti-inflammatory responses to n-3 PUFAs are not fully understood; furthermore, many effects from n-3 PUFA treatment are not well characterized in humans. Of interest is the role of DNA methylation (DNAm) in mediating the effects of n-3 PUFAs on inflammation.

**Objective:** We aimed to characterize the effects of n-3 PUFA treatment on DNAm in inflammation-related signaling pathways in PBMCs of women at high risk of breast cancer

**Methods:** PBMCs of women at high risk of breast cancer were obtained at 0 and 6 months of n-3 PUFA treatment in a previously reported dose finding trial (n=10 matched pairs in the 5 g/day EPA+DHA dose arm).[53] DNA methylation of PBMCs were assayed using reduced representation bisulfite sequencing to obtain genome-wide methylation profiles on a single nucleotide level. Analyses were performed to investigate the effects of n-3 PUFA treatment on DNAm both genome-wide and within a set of candidate genes.

**Results:** A large number of differentially methylated CpGs (DMCs) in gene promoters (24,842 DMCs in 5507 genes) showed significant enrichment for hypermethylation in both the candidate gene and genome-wide analyses. Using these DNAm changes, pathway analysis identified significantly hypermethylated signaling networks after n-3 PUFA treatment, such as the Toll-like Receptor pathway. Based on analyses of data per individual, DNAm changes from n-3 PUFA treatment appear highly variable between study participants.

**Conclusions:** Dietary n-3 PUFA supplementation for six months is associated with DNAm changes in PBMCs with potential for anti-inflammatory effects. PBMC DNAm profiles may offer a novel means of assessing individual response to n-3 PUFAs. This observation warrants further investigation in future n-3 PUFA intervention studies.

## Introduction

Dietary n-3 polyunsaturated fatty acids (PUFAs) have health benefits for a wide range of diseases in both preclinical and clinical studies, particularly for conditions with underlying chronic inflammation such as cardiovascular disease, rheumatoid arthritis, dementia, and cancer.[10, 46] In humans, levels of eicosapentaenoic acid (EPA) and docosahexaenoic acid (DHA) are largely dependent on consumption of foods or supplements enriched in these fatty acids such as fatty fish or fish oils, given the inefficient conversion of plant-derived alpha linolenic acid to long chain n-3 PUFAs.[9] Intervention studies of dietary n-3 PUFAs/fish oil indicate capacity for increased EPA, DHA, and n-3:n-6 PUFA ratio in plasma, adipose tissue, and circulating cells such as erythrocytes and peripheral blood mononuclear cells (PBMCs).

Clinical investigations of dietary EPA and DHA have demonstrated anti-inflammatory effects of n-3 PUFA supplementation by assessing responses of PBMCs. Previous studies involving healthy volunteers showed that fish oil supplement reduced the production of cytokines including interleukin 2 (IL-2), interleukin 1 (IL-1), and tumor necrosis factor alpha (TNF*α*) by mononuclear cells.[13, 14, 17] Gene expression studies of PBMCs from healthy subjects taking fish oil (775 mg/d EPA) and borage oil (831 mg/d gamma linolenic acid) supplements for four weeks demonstrated decreased expression of PI3K*α* and PI3K*γ*, key mediators of pro-inflammatory signal transduction, and cytokines (IL-1, IL-10, IL-23) via quantitative RT-PCR.[51] In a study of older adults, treatment with 1.8 g/day EPA+DHA for six months resulted in PBMCs with decreased pro-inflammatory gene expression involving the interleukin, MAP kinase, NF-*κ*B, and Toll-like receptor signaling pathways.[8] DHA-rich n-3 PUFA supplements (1.7 g/d DHA, 0.6 g/d EPA) versus corn oil placebo for 6 months in older adults with Alzheimer’s disease led to decreased production of IL-6, IL-1*β*, and granulocyte colony stimulating factor by PBMCs stimulated ex vivo by lipopolysaccharide (LPS), without decreases in TNF*α*, IL-10.[49] Corresponding analyses of PBMC gene expression showed changes consistent with an anti-inflammatory effect, although without correlation to the cytokines and growth factors induced by LPS.[50] The specific cytokines affected may relate to the dose, duration, and composition of n-3 PUFA supplements in addition to patient-related factors.

Epigenetic regulation of gene expression may mediate, at least in part, the anti-inflammatory effects of dietary n-3 PUFAs. DNAm is one mechanism by which pro-inflammatory genes are silenced, as shown in prior reports of epigenetic regulation of inflammation in a wide range of cell types including PBMCs,[6, 44] CD4+ lymphocytes,[35] and breast cancer cell lines.[8, 19] Regarding the role of dietary n-3 PUFAs in modulating the epigenome, a cross-sectional study of DNAm in PBMCs of Yup’ik Alaskan Native Americans at the lowest and highest range of long chain marine n-3 PUFA intake showed potential impact of DNAm on anti-inflammatory pathway genes.[5] Modulating effects of n-3 PUFAs on DNAm that affect inflammatory and metabolic signaling pathways are also evident in short term intervention trials of n-3 PUFAs.[12, 21, 47]

To further elucidate the effects of dietary n-3 PUFAs on epigenetic regulation of inflammation-related signal transduction, we examined DNAm changes in PBMCs of women at high risk of breast cancer following six months of n-3 PUFA supplementation. We utilized reduced representation bisulfite sequencing (RRBS) to analyze DNAm on a genome-wide level with single base-pair resolution and coverage of many more sites than array-based methods.

## Methods

### PBMC

PBMCs were isolated via ficoll-hypaque separation from peripheral blood collected in acid citrate dextrose (ACD) blood tubes as part of a study of n-3 PUFAs in women at high risk of breast cancer; the main results of the trial were previously reported.[53] In brief, 48 women at high risk of breast cancer were randomly assigned to one of four daily doses of n-3 PUFAs (0.84, 2.52, 5.04, or 7.56 g/d of EPA+DHA) for 6 months of treatment. The study was conducted with the approval of the Institutional Review Board of The Ohio State University and in accordance with ethical standards of the 1975 Helsinki Declaration and its later amendments. PBMCs were processed within 4 hours of collection and cryopreserved in liquid nitrogen until analysis. In the present study, we utilized PBMCs obtained at 0 and 6 months of 5.04 g/d EPA+DHA from the 10 of 12 sample sets with highest increase in DHA/EPA at 6 months. This dose is utilized in an ongoing phase II trial (clinicaltrials.gov ID NCT02295059).

### DNAm Data Generation, Processing, and Quantification

DNAm data was generated for all samples using the RRBS method.[31] Prior to library generation, DNA was extracted using QIAamp DNA mini kit from QIAGEN. Sequencing libraries were created using the NEXTflex Bisulfite Library Prep Kit for Illumina Sequencing from BIOO following the manufacturer’s protocol. All samples were sequenced using 100bp paired-end technology. Sequenced reads were first trimmed for both adapter sequences and low quality using TrimGalore v0.4.0 (a wrapper for Cutadapt) which removed any detected Illumina adapters or bases with Phred quality scores below 20 from the 3’ end of each read.[33] Next, reads were aligned using Bismark v0.17.0 to the human genome (version GRCh38).[40] Bismark alignment was performed using Bowtie 2 and the minimum alignment scoring function set to “L,-0.6,-0.6”. Quality was assessed using FastQC v0.10.1, trimming reports, and alignment reports to ensure low adapter, low quality trimming rates, low incomplete conversion, and acceptable unique alignment rates for each sample. Additional reads were sequenced until each sample had 30 million reads that passed the quality filter. After passing our quality assessment, DNAm was quantified as the percent of CpG sites that are methylated in the sample using Bismark’s “bismark methylation extractor” function. Additional processing of DNAm quantification (i.e. filtering CpGs based on read depth, extracting coverage, generating coverage plots, etc.) was performed by custom scripts written in R v3.3.2 and Python v2.7.8.

### Power analysis and Sample Size Determination

To determine how many samples should be included in this study, we performed a power analysis based on detecting average DNAm differences at CpG loci of 5% in PBMCs. The variance for PBMC DNAm was determined using publicly available data. Control PBMC samples assayed for DNAm using the Illumina Infinium 450k array were used to determine the standard deviation (SD) in DNAm at CpGs as a function of DNAm (GEO series GSE57107). A fit of the SD values produced a maximum value at 50% DNAm (max of fit SD values = 0.30; data not shown). For the power analysis, a more conservative SD of 0.65 which corresponds to the 95th percentile of all CpGs was used. The power analysis was conducted using an FDR corrected alpha = 0.05 to account for multiple testing of the CpGs in the promoter region of our candidate genes. We found that 20 total samples (10 baseline and 10 after treatment) produced sufficient statistical power for detecting a 5% difference in average DNAm of 96%. Based on this analysis, we selected 10 paired samples (10 baseline and 10 after six months n-3 PUFA treatment) with the most consistent increase in EPA and DHA of the 12 participants in the 6 capsule/day dosing arm of the n-3 PUFA phase I clinical trial.[53] The variation in our DNAm data, which was assayed using the RRBS method, was lower than in the public data used for the power analysis (Supplemental Figure 4).

### Summarizing DNAm Genome-wide

Global DNAm levels were determined using the total number of unconverted (methylated) cytosines at CpG sites divided by the total coverage at CpG sites genome wide. Only CpGs with at least 5 reads coverage in at least 3 samples in both treatment groups were included in the global DNAm average. The treatment group averages for all CpGs that achieved the coverage and sample threshold were plotted to create experiment wide DNAm scatter plots. All plotted CpGs were fitted using orthogonal regression (ODR function from the SciPy library in Python) with the standard deviation used as the error for each CpG (SciPy.org). We selected orthogonal regression, as opposed to the more traditional linear regression, as it provides the most accurate estimation of the error in the case of our null hypothesis of no DNAm change from n-3 PUFA treatment (illustrated by the y=x line). Linear regression assumes that only one variable produces the observed error for each measurement whereas orthogonal regression assumes that both variables contribute to the observed error.

### Genomic Feature Summaries

DNAm for annotated genomic features were summarized differently. First, all CpGs within a region that had at least five reads coverage were averaged to produce an average DNAm for each region. Next, the averages for all regions that comprise a genomic feature were again averaged into one DNAm value that was used to represent the DNAm for that genomic feature. Feature summaries were performed on the genomic features where DNAm has been implicated as playing a role in regulating gene expression. Therefore, CpG islands, two definitions of the promoter region, CpG islands located in the distal promoter, gene bodies, and the 1st coding exon of each gene were interrogated for DNAm changes. The two promoter regions were defined to create a long-range and a short-range promoter (10kb upstream, 1kb downstream and 1kb upstream, 1kb downstream, respectively, of the transcriptional start site [TSS]).

### Candidate Gene List Development

All GDAC Firehose analysis results and data were downloaded to access their 8,586 gene DNAm table (Broad Institute TCGA Genome Data Analysis Center [GDAC] 2012). The DNAm table provided data in the format of 450k Infinium array beta-values for each of the 8,586 most varying probes which were associated with gene symbols. Pathway membership for each gene was obtained by querying against Gene Ontology, KEGG, and Reactome using each site’s API.[26, 27, 39] Genes involved in inflammation related pathways were kept resulting in 89 inflammation related genes that are highly variable in breast cancer. An additional 63 genes were identified by performing literature searches in June of 2015 and again in May of 2016 using combinations of the main keywords in both PubMed and Google Scholar: n-3 PUFA, fatty acids, n-3 PUFA, inflammation, DNA methylation, PBMC, cancer, breast cancer, prevention, biomarker. These 63 genes and their references are provided in Supplemental Table 2. The genes that resulted from the literature search were all reported to play a role in or have been affected (DNAm or expression levels) by fatty acid metabolism, fatty acid related inflammation, or breast cancer inflammation. The resulting 152 genes (Supplemental Table 2) are the candidate genes.

Unsupervised clustering was performed using the same non-negative matrix factorization (NMF) method used in the original GDAC analysis, but was implemented in R using the NMF package (v0.20.6) for analysis convenience[29] (Supplemental Figure 1A). Analysis in R was successful in reproducing sample membership of clusters reported by GDAC. To perform NMF on candidate genes, the full 450k Infinium Array data was downloaded from TCGA for the entire breast cancer cohort so that DNAm beta values could be obtained. Metadata (sample and experiment numbers as well as clinical classification) were downloaded so that sample IDs could be converted and matched. Using only the samples included in the original GDAC analysis and selecting the most varying probe for each candidate gene, the 152 candidate gene beta-value table was constructed. NMF was performed and quality measures calculated for 2-10 clusters. As in the GDAC analysis, the quality measures cophenetic similarity, dispersion, and silhouette coefficients all indicated that six clusters resulted in the best separation (Supplemental Figure 1C). Molecular markers ER, PR, and HER2, Triple Negative status, and PAM50 subtype, were all used to determine if the six resultant sample clusters corresponded to clinically relevant classifications. Both the original 8,586 GDAC clusters and the candidate gene clusters were annotated with clinical classification to observe associations. However, as with the original clustering, clustering did not stratify samples based on any single or combination of molecular markers included in the metadata. Although clustering could not be explained by molecular markers, we did observe that both the original GDAC cluster and our clustering showed a general separation between luminal and basal-like breast cancers. For both the GDAC gene list and our candidate gene list, the majority of the basal-like samples were present in a single cluster (Supplemental Figure 1 B&D). The stratification of these subtypes supports the observed differential expression in breast cancer marker genes that was also reported in the original GDAC analysis. Additionally, the fact that our candidate gene list performs similarly implies that our greatly reduced gene list is also able to discern differences in the genes responsible for resulting in different breast cancer subtypes.

### DMC Analysis and Association with Gene Promoters

DMCs were determined between treatment groups using MethylKit v1.2.0.20 All CpGs with 10 reads coverage in at least 16 samples were input into MethylKit resulting in a total of 962,093 CpGs being tested (Figure 2A). A DMC had a SLIM multiple test corrected p-value less than 0.05 and at least a 5% average DNAm change between treatment groups. DMCs in candidate genes were found by intersecting all DMCs with the promoters and promoter associated CpG islands of the candidate genes. Promoter DMCs were identified as any DMC that was in a region 10k base-pair (bp) upstream and 1k bp downstream of transcriptional start sites of RefSeq genes (hg38). To ensure that DMCs were associated with n-3 PUFA treatment effect and not explained by any confounders, we performed linear regression and correlation analysis for each DMC. Using linear regression, DNAm at DMCs of all samples were compared to both the fatty acid levels and estimated cell type composition as determined by methylCC [20]; DMCs where the resulting fit had an FDR p-value ≤ 0.1 with any confounding variable were removed. The maximum R^2^ of the DMCs that passed this filtering process and were used in subsequent analyses was 0.58. Additionally, we used a paired t-test to test for changes in the estimated fraction after n-3 PUFA treatment for each cell type present (i.e. granulocyte, CD4+, CD8+, B cells, monocytes, natural killer cells); no significant changes were detected (data not shown).

**Figure 1.**
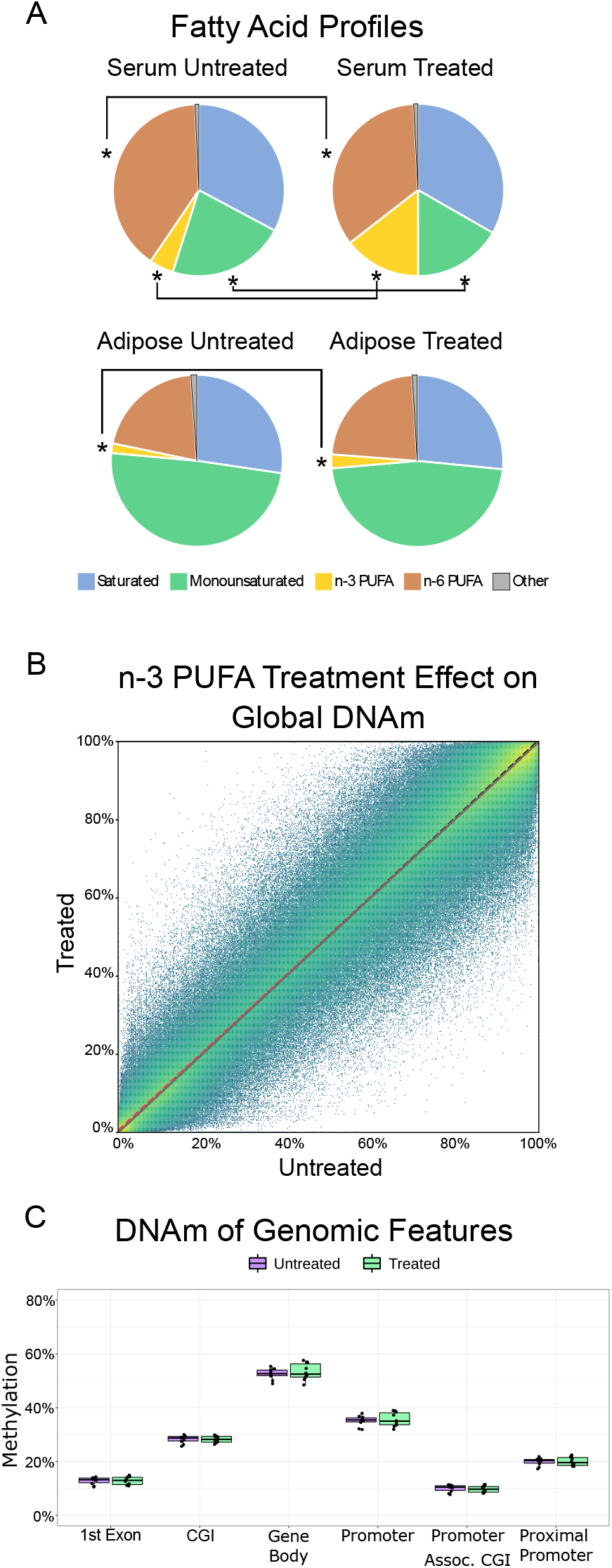
Effects of n-3 PUFA treatment on fatty acid profiles and averaged DNAm. A) Average fatty acid content before (left; n=10) and after (right; n=10) 6 months of n-3 PUFA treatment. The fatty acid composition was measured in both serum (top) and adipose tissue (bottom) as previously detailed.[53] Asterisks indicate statistically significant changes from n-3 PUFA treatment (FDR p-value ≤ 0.05). B) Global DNAm represented by all CpGs that had 5 reads in at least 3 samples. The black dotted line is the y=x line which indicates no change in average DNAm. The color (blue to yellow) indicates the number of CpGs represented by each point. Data were fitted using orthogonal regression (white dashed line). The fit showed no global effect from n-3 PUFA treatment. C) DNAm averaged at 0 and 6 months for six genomic features. Genes are RefSeq GRCh38 annotation, and genomic features are defined as follows: 1) CpG Islands (CGI) – UCSC genome browser CpG island track; 2) Promoter associated CGI –CGI that fall within 10kb up-/1kb downstream from gene transcriptional start sites (TSS); 3) Promoter – region defined as 10kb up-/1kb downstream from TSS; 4) Proximal promoter – promoter region defined as 1kb up-/1kb downstream of TSS; 5) first exon; 6) Gene body – entire gene coding region for all transcripts. There were no significant treatment effects (mean ± standard error) in these summary features by averaged DNAm.

**Figure 2:**
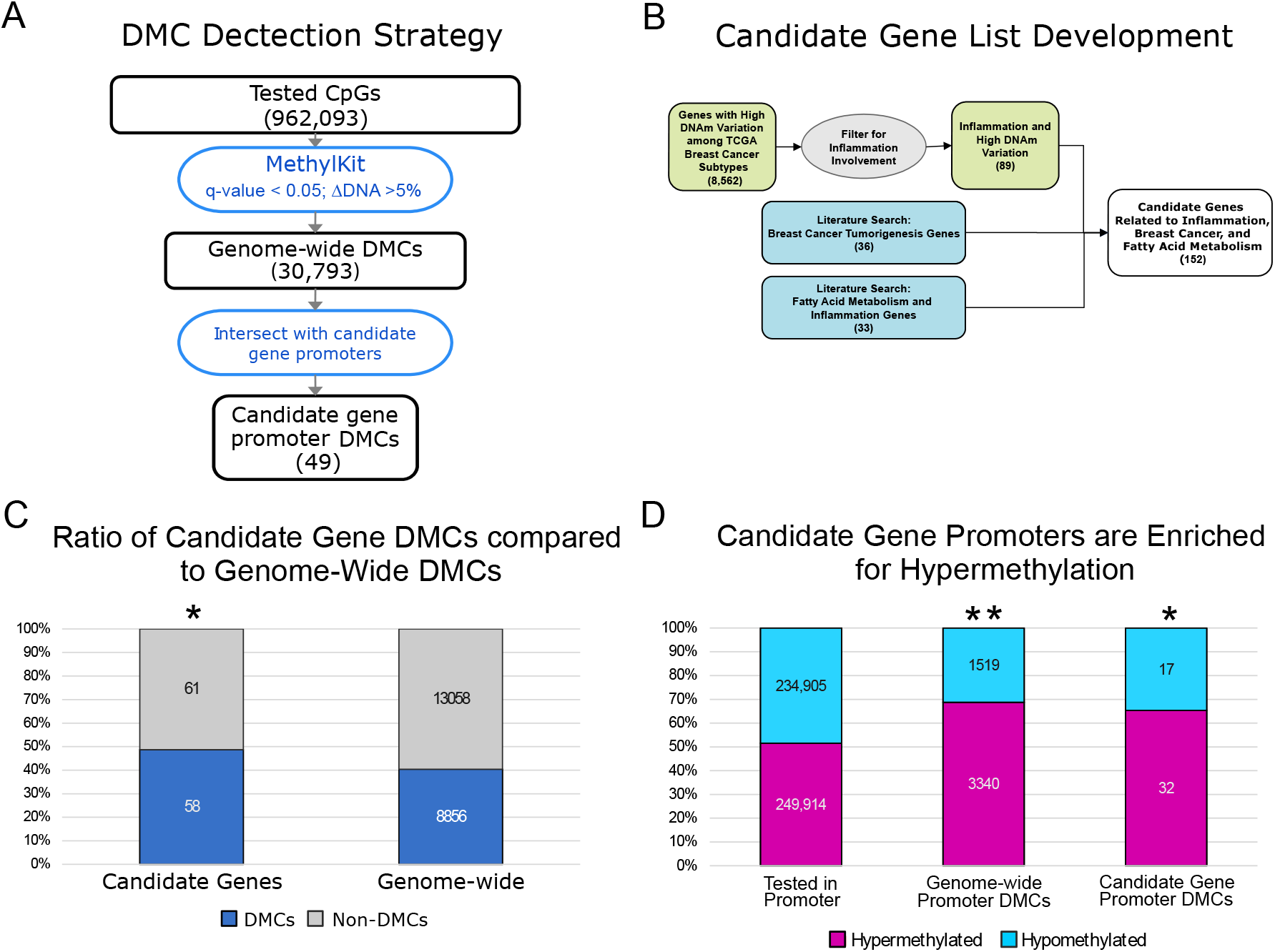
Differential DNAm in a set of Candidate Genes after n-3 PUFA Treatment. A) The flow chart depicts the processing involved in identifying differentially methylated CpGs (DMCs). All CpGs with at least 5 reads coverage in 80% of the samples (16/20 samples) were included in the analysis (962,093 CpGs). DMC analysis was performed using MethylKit. CpGs that had a multiple test corrected p-value (q-value) less than 0.05 and a change in DNAm (ΔDNAm) ≥ 5% were considered DMCs. Candidate gene promoter DMCs were identified as those that overlap the genomic coordinates of the candidate gene promoters (defined as 10kb up-, 1kb downstream of the TSS). B) A flow chart for the development of the candidate gene list. The 152 candidate genes were based on either 1) genes with highly variable DNAm in the TCGA breast cancer cohort that involve inflammation or 2) literature review for inflammation related DNAm processes, DNAm and fatty acids or breast cancer development. C) The candidate genes (left bar) were enriched in DMCs when compared to all genes (right bar; hypergeometric test p-value = 0.026). D) Genome-wide promoters and candidate gene promoters were enriched in hypermethylation after n-3 PUFA treatment. The hypergeometric test was used to compare the ratio of hyper- to hypo-methylated genes in all tested gene promoters (left bar) to both the genome-wide promoter DMCs (** middle bar; p-value ≤ 0.001) and the candidate gene promoters (* right bar; p-value = 0.018).

### DMC Pathway Analysis and Directional Enrichment

Genome wide promoter DMCs were analyzed for enrichment of inflammation related pathways to show the involvement of inflammation in PUFA treatment. DMCs were input into IPA[34] and the Enrichr[1] web tool to obtain results from Biocarta, KEGG, NCI-Nature, Panther, Reactome, and WikiPathways.[28, 36, 38, 39, 42, 42, 45] All significantly enriched pathways (adjusted p-value ≤ 0.05) were extracted. Inflammation related pathways were identified based on processes previously reported to be involved in inflammation.[1, 28, 38, 42, 45, 45, 48]

### Enrichment of Pathways for Directional DNAm Change

Candidate gene promoter DMCs were used to determine which inflammation-related WikiPathways were enriched in n-3 PUFA treatment DNAm changes.[42, 45] Directional enrichment was determined using the genome-wide promoter associated DMCs instead of the candidate gene promoter DMCs that were used for general enrichment.[32] The DNAm change from n-3 PUFA treatment was determined for each gene in each pathway using all DMCs in the promoter region. We tested for an over-representation of either hyper- or hypomethylated DMCs in each identified pathway that included at least 8 DMCs. Only genes that had a DMC in the promoter region were used to determine directional DNAm enrichment. Because the genome-wide DMCs had a strong trend toward hypermethylation (7,129 hyper- vs 3,174 hypomethylated DMCs), the hypergeometric test was used to determine if the ratio of hyper- to hypomethylated DMCs in each pathway was significant given the genome-wide DMC background. Pathways were considered to be significantly enriched if the hypergeometric test produced a q-value ≤ 0.05 (FDR corrected p-value).

### Correlating DNAm with Fatty Acids, Serum Biomarkers, and Biometric Data

For each DNAm measure and each patient data type (i.e. total fatty acid, serum biomarkers, etc.), DNAm was correlated with the patient data using linear regression. The resulting linear model was assessed by producing an R^2^ value and multiple test corrected (FDR) p-value to control for testing multiple DNAm measures and the multiple parameters included in the data type. Using the total fatty acids data type as an example, all total fatty acid measures (n-3 PUFA, n-6 PUFA, monounsaturated, saturated) were compared to DNAm of all six genomic features and the resulting p-values were corrected for 24 comparisons. These correlations were performed for the baseline values, the change after n-3 PUFA treatment and the baseline vs n-3 PUFA change. This analysis used DNAm measures of global DNAm, genomic feature summaries, and candidate gene promoter DMCs. Each DNAm measure was compared to the patient data for total fatty acids, individual fatty acids, serum inflammation markers (Adiponectin, Leptin, high-sensitivity C-reactive protein, IL-6, and TNF*α*), and biometric data (age, weight, BMI, waist, waist hip circumference).

## Results

### n-3 PUFA treatment does not alter average genome-wide DNA methylation patterns

We previously reported the effects of four different doses of n-3 PUFAs on fatty acid profiles of women at high risk of breast cancer following six months of treatment.[53] For this PBMC DNAm substudy of 10 of 12 participants from the 6 capsule/day arm, analysis of serum and breast adipose fatty acids yielded similar results to those previously reported.[53] The 10 women had an average age of 51 ± 6.8 years, weight of 164.5 ± 25.8 lbs., BMI of 27.1 ± 5.0, waist of 84.7 ± 9.3 cm, and waist hip circumference ratio of 0.80 ± 0.06 (mean ± SD). Averaged pre- and post-treatment serum fatty acid profiles showed changes in the overall distribution of fatty acids, with significant increases in total n-3 PUFAs and decreases in total monounsaturated fatty acids and n-6 PUFAs after 6 months of study treatment (Figure 1A; Supplemental Table 1). Fatty acid profiles in adipose tissue also showed significantly higher n-3 PUFA content at six months compared to baseline (Figure 1A).

To assess whether n-3 PUFA treatment leading to increased serum and adipose EPA, DHA resulted in a global remodeling of DNAm patterns, we calculated the effect on average DNAm before and after treatment (n=10 matched pairs). For a genome-wide view of DNAm, the average DNAm values of all pass-filter CpGs (≥5 reads coverage in at least 80% of samples) were plotted and the line of best fit was used to determine whether n-3 PUFAs caused a global change in DNAm (see Methods). The resulting fit, with an R^2^ of 0.999, indicates that DNAm averaged before and after treatment is unchanged at most CpG sites (Figure 1B). The average DNAm change genome-wide was only 0.7% (50.5% pre- and 51.2% post-treatment, t-test p=not significant). We also determined the effect of dietary n-3 PUFAs on DNAm of specific regions of the genome by evaluating the change in DNAm in six annotated genomic features that are associated with gene regulation (e.g. gene promoters and CpG Islands). In these selected genomic features, average DNAm did not change after n-3 PUFA treatment (Figure 1C).

### n-3 PUFAs resulted in promoter hypermethylation as assessed by genome-wide and candidate gene CpG-level analyses

To determine how n-3 PUFA effects on DNAm could potentially regulate gene expression, we investigated the average DNAm effects at CpG loci. A genome-wide differentially methylated CpG (DMC) analysis was conducted with MethylKit to compare the DNAm between 0 and 6 months.[2] All CpGs in the genome with sufficient read coverage were tested for changes in the average DNAm (see Methods). To limit the detected DNAm changes to only those caused by the n-3 PUFA treatment, DMCs with DNAm changes significantly correlated with estimated cell type were removed (180 DMCs removed; see Methods).[20] This analysis at CpG loci identified a total of 30,793 DMCs following n-3 PUFA treatment (DNAm change ≥ 5% and multiple test corrected p-value≤0.05; Figure 2A). To reduce the number of DMCs and focus on those more likely to regulate gene expression [7, 24], the DMCs in the promoter region (10kb up- and 1kb down-stream from the TSS) were examined. However, this filtering still resulted in 10,261 DMCs located in 5,491 gene promoters.

Instead of performing a genome-wide analysis to investigate all DNAm-mediate effects from n-3 PUFA treatment, we focused the analysis based on our hypothesis that n-3 PUFA treatment affects inflammation and breast cancer pathways. Before conducting experiments, we developed a set of candidate genes based on two criteria: 1) genes involved in inflammation that showed high levels of variation and stratified the breast cancer subtypes in the Cancer Genome Atlas (TCGA) breast cancer data set (89 genes; GDAC Analysis 2012; see Methods for more details) and 2) genes associated with breast cancer (36 genes) and fatty acid induced effects (33 genes) (Figure 2B; Supplemental Table 2). Using the resulting candidate genes (Supplemental Table 2), we demonstrated a similar stratification of the TCGA data when comparing the original 8,562 genes to our reduced set of 152 candidate genes (Supplemental Figure 1). From the genome-wide promoter DMCs, candidate gene DMCs were identified by selecting only those DMCs located in a candidate gene promoter region (Figure 2A). This approach both focused the analysis to our hypothesis and used a more stringent genome-wide multiple test correction to reduce false positives. The full genome-wide promoter DMCs were used later in our analysis to predict the effects of n-3 PUFA treatment.

From the 152 candidate genes, we identified 49 candidate genes that contained at least one DMC (compared to 5,491 genome-wide gene promoters). Comparing the proportion of candidate gene DMCs to genome-wide DMCs, the candidate genes were significantly enriched in n-3 PUFA associated DMCs (hypergeometric test qval = 0.026; Figure 2C-D). Both the genome-wide and candidate gene promoters were significantly enriched in hypermethylation after n-3 PUFA treatment, with 68% hypermethylated genome-wide promoters and 65% hypermethylated candidate gene promoters compared to 52% hypermethylation in all gene promoters (Figure 2D).

### Pathway analysis identifies potential epigenetic mechanisms of n-3 PUFAs

To focus on those pathways of interest for n-3 PUFA effects, we then used the candidate gene promoter DMCs to perform pathway analysis. We identified 31 pathways enriched for candidate gene promoter DMCs (adjusted p-value ≤ 0.05; Supplemental Table 4). To determine which pathways were more likely to be up- or down-regulated by the n-3 PUFA-induced DNAm changes, we utilized the genome-wide promoter DMCs. The DNAm change in each gene promoter DMC was used to identify the pathways where n-3 PUFA treatment caused concerted treatment effects (all DMCs showing either hyper- or hypomethylation). Enrichment in directional DNAm was determined using the hypergeometric test in all pathways with at least 8 genome-wide promoter DMCs (minimum number of DMCs to achieve hypergeometric p-value ≤ 0.05).[43] This test yielded three pathways significantly enriched in n-3 PUFA associated DNA hypermethylation: the Focal Adhesion, Toll-like Receptor (TLR), and Leptin signaling pathways (Figure 3A). Of these pathways, the Focal Adhesion and TLR pathways had both enrichment for DNA hypermethylation and biologically relevant localization of DMCs (Supplemental Figures 2-3). By comparing the DMCs detected in these pathways (Figure 3B) and the genes that comprise each pathway (Figure 3C), we noted only two shared DMCs and two shared signal transduction pathways. The localization of DMCs within each pathway has potential biological relevance because of the canonical relationship between increased DNAm in gene promoters and gene silencing. Notably, the hypermethylated DMCs in the Focal Adhesion pathway are located downstream of cytokine, chemokine, and hormone signaling (Supplemental Figure 2). The TLR pathway has hypermethylated DMCs downstream of lipopolysaccharide (LPS) receptors and upstream of pro-inflammatory cytokines (Supplemental Figure 3). Taken together, these pathways represent two distinct mechanisms by which n-3 PUFAs may modulate inflammation through DNAm changes.

**Figure 3:**
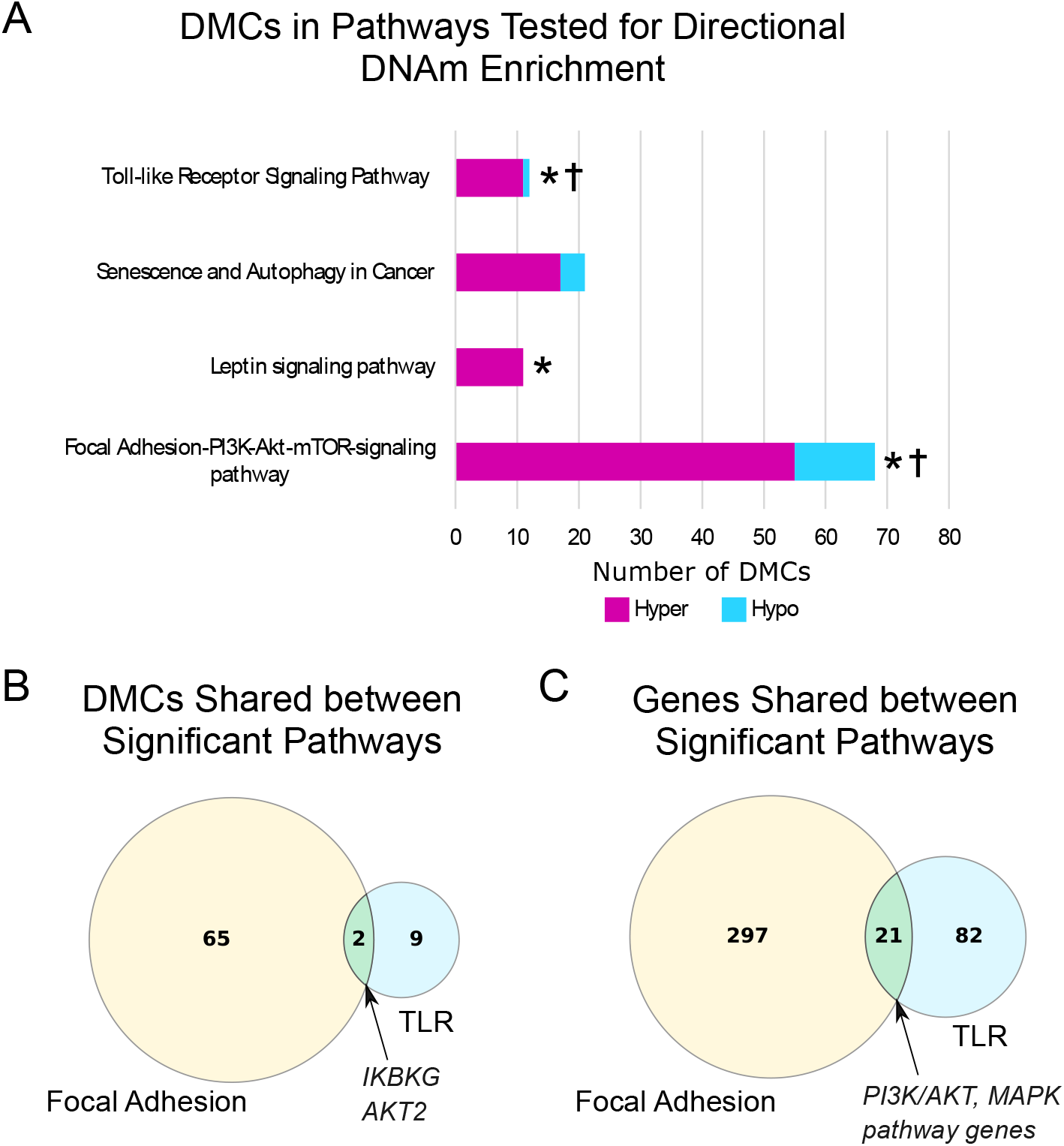
Directional Enrichment of DNAm Changes in Candidate Gene Associated Pathways. Inflammation-related pathways were determined using DMCs located in candidate gene promoter regions. A) The hypergeometric test was used to compare the number of hyper- or hypomethylated DMCs observed in each pathway to the distribution of DMCs in promoter regions genome-wide (7,129 hyper- and 3,174 hypomethylated DMCs). Asterisks indicate hypergeometric FDR p-value ≤ 0.05. Daggers indicate that hypermethylated DMCs are localized in biologically relevant region(s) of each pathway. Of the four pathways that were tested for directional enrichment, three had significant overrepresentation of hypermethylated DMCs (FDR p-value ≤ 0.05; indicated by stars). Two pathways were both enriched in DMCs and biologically relevant localization of DMCs. B) Comparison of the promoter DMCs used to determine enrichment in the Focal Adhesion and TLR pathways shows the shared genes. Comparison of all genes included in the Focal Adhesion and TLR pathways shows an overlap of 21 genes. The shared genes represent two shared signaling cascades in both pathways.

### Exploratory analyses of variability in PBMC DNAm after n-3 PUFA Treatment

Although average DNAm did not change following n-3 PUFA treatment in any of the six genomic regions analyzed (Figure 1C), we observed highly variable DNAm patterns when comparing DNAm change in the genomic features for each sample pair from 0 to 6 months (Figure 4A). Similarly, n-3 PUFAs did not affect average global DNAm (Figure 1B), but the change per individual was highly variable (Figure 4B; mean = 0.7%; minimum= −10.5%; maximum = 12.1%). The individual fatty acid profiles did not show a similar variability between the participants (Figure 4C).

**Figure 4:**
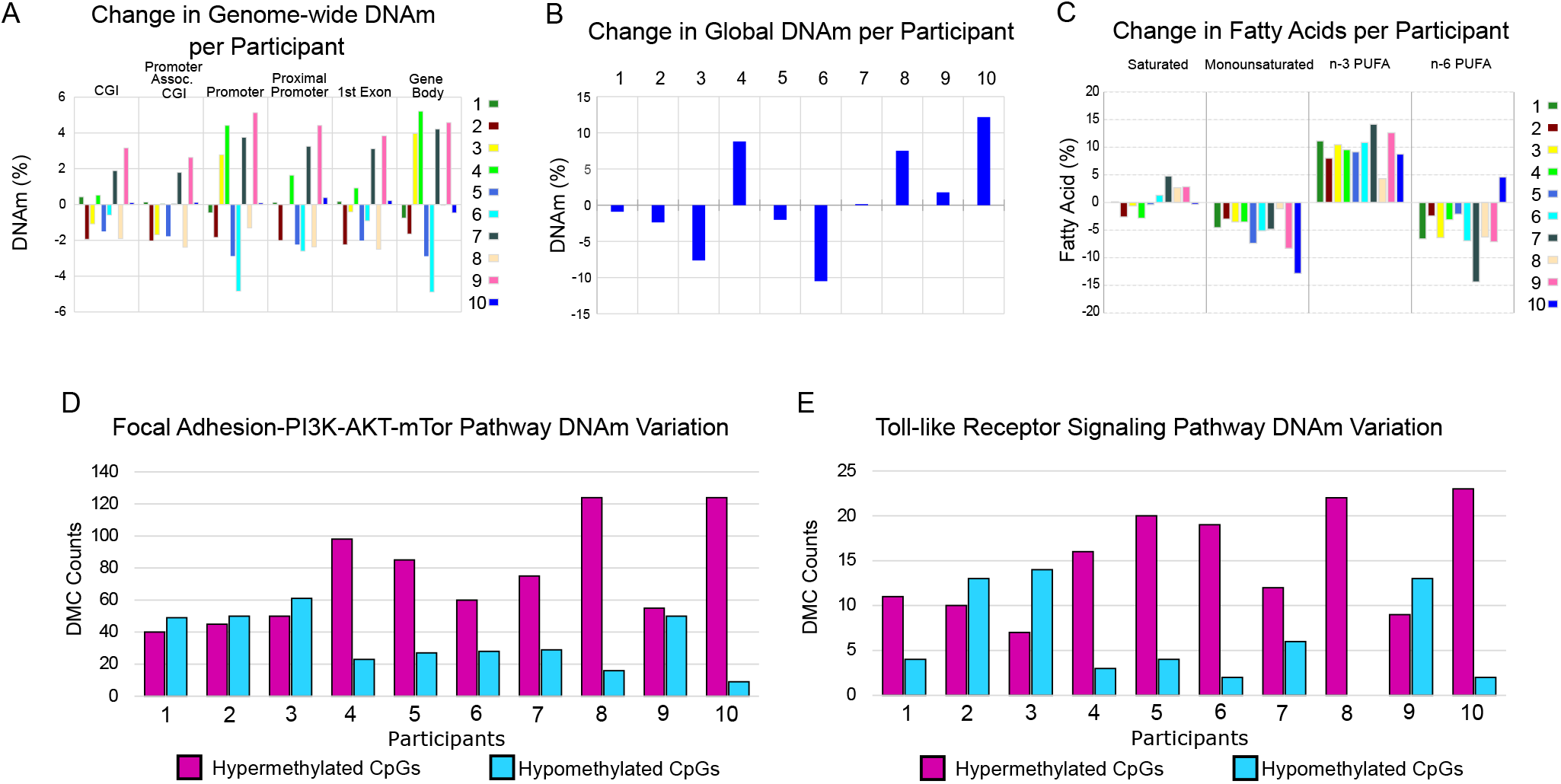
n-3 PUFA Treatment Effects show High Variability Between Samples. A) n-3 PUFA treatment produces highly variable effects on genome-wide DNAm between participants in gene associated features. Participants are labeled 1 through 10. The DNAm was averaged for each of the genomic features for each participant at 0 and 6 months of n-3 PUFA treatment. The change in DNAm from n-3 PUFA treatment (treated minus untreated) per individual is shown. B) The change in global DNAm also showed variable treatment effects between women. The average DNAm values from all CpGs with at least 5 reads coverage were used to determine global DNAm for each participant at 0 and 6 months. C) Following n-3 PUFA treatment, the serum fatty acid profiles of all participants showed increases in n-3 PUFAs and decreases in monounsaturated fatty acids. Following n-3 PUFA treatment, participants differed in directional change of saturated fatty acids and n-6 PUFAs. For each fatty acid measure, the change after n-3 PUFA treatment is shown (treated minus untreated) for each sample pair. D-E) Variable n-3 PUFA treatment effects at the CpG level were assessed in the Focal Adhesion and TLR pathways. For each study participant, the number of hyper- and hypo-methylated CpGs were counted for each of the DMCs detected in the two pathways. CpGs were counted only when the magnitude of the DNAm change was greater than 5% so that the variability was not overrepresented by small changes. Based on the average effects, more hypermethylated CpGs would be expected; however, several participants had PBMCs with a higher number of hypomethylated CpGs. In four participants, PBMCs showed relatively more CpG hypomethylation than hypermethylation in at least one pathway at 6 months of n-3 PUFAs; for participants 2 and 3, there was greater CpG hypomethylation in both pathways.

To investigate the basis for the observed individual vs average differences in DNAm in the 10 participants, we tested for correlations of DNAm with possible confounders such as BMI, age, waist, serum biomarkers (i.e. adiponectin, leptin, high-sensitivity C-reactive protein, IL-6, and TNF*α*), fatty acid profiles (EPA, DHA, n-3 and n-6 PUFA, poly- and mono-unsaturated fatty acids), and estimated cell type composition (natural killer, monocytes, B-cells, CD8+ T-cells, CD4+ T-cells, granulocytes) using both baseline values and change from treatment. The analyses did not reveal significant correlations between potential confounders and DNAm (global or genomic features; see Methods for further details; data not shown).

Given the observed variability in global DNAm between participants, we assessed whether the Focal Adhesion and TLR pathways, as identified by pathway analysis of statistically significant differences in average DNAm, also had variable DNAm change between participants at the CpG level. Some women showed PBMC DNAm changes in the opposite direction when compared to the average. To illustrate this variability, we counted the number of hypo- vs hyper-methylated CpGs for each study participant for all DMCs in each of the pathways; this showed that some participants had more hypomethylated CpGs (Figure 4D-E; CpGs with DNAm change ≥ 5%).

## Discussion

In this work, we demonstrate that n-3 PUFA supplementation elicits a strong, locus-specific effect on DNAm of PBMCs in a study cohort of women at high risk of breast cancer. While average genome-wide CpG methylation was essentially unchanged in PBMCs after six months of n-3 PUFA supplementation, analysis of DNAm changes at CpGs (DMCs) revealed marked enrichment of hypermethylation in the promoter regions of candidate genes selected for potential for epigenetic regulation in inflammation and breast cancer. Using the DNAm changes in gene promoters of the candidate genes, pathway analysis identified at least two pathways that might in part mediate n-3 PUFA anti-inflammatory effects.

Our DNAm data also indicate a highly variable individual response to n-3 PUFA treatment in matched pairs of DNAm data at 0 and 6 months. Exploration of the genome-wide DNAm changes of each sample pair revealed differing and even opposing DNAm effects from n-3 PUFA treatment at every scale in the genome: global DNAm (Figure 4B), gene associated genomic features (Figure 4A), and individual CpGs (Figure 4D-E). Using linear regression methods, variability in DNAm patterns did not correlate with demographic or anthropometric features with known effects on DNAm such as age, menopausal status, and central obesity.[3, 11] Prior studies have noted individual variability in response to dietary n-3 PUFAs that relates to baseline parameters (e.g. fatty acid levels, BMI)[53] or genotype (e.g. variants in COX1, IL-6, FADS1, and FADS2),[30, 41, 47, 48] with potential for differential treatment effects. With the lack of consensus regarding clinical use of n-3 PUFAs, as for depression, cardiovascular disease, and cancer,[15, 22, 23, 52] PBMC DNAm profile might provide another measure of individual response to n-3 PUFAs. However, as our initial sample size estimates were based on the analysis of averaged DNAm, these observations of variability in individual DNAm response fall outside the purview of the current study.

Our findings on n-3 PUFA effects on averaged DNAm in PBMCs are largely consistent with prior reports of average gene expression changes after n-3 PUFA treatment. We observed both direct methylation of pro-inflammatory genes and hypermethylation of multiple mediators in pro-inflammatory pathways that have previously been reported to show changes in gene expression. The pro-inflammatory genes IRAK1, CXCL16, ALOX5, MAN2A1, HSD17B11 were hypermethylated in our study cohort, which corroborates decreases reported in previous n-3 PUFA studies.[8, 49] We also observed hypermethylation upstream of the pro-inflammatory genes TNF, IL1B, IL6, IL12A which corroborate the findings of others.[8, 18, 19, 37] reported significant downregulation of the TLR and the MAP kinase signaling pathways, which we also identified as significantly enriched in hypermethylated DMCs.[8] In one of the most comprehensive reports of n-3 PUFA’s effects on DNAm, Tremblay et al. used the 450k Infinium array to demonstrate that 6 weeks of n-3 PUFA supplementation led to hypermethylated CpGs and changes in pathways that included the inflammation-related related TLR and PI3K pathways.[47] Thus, the results of our analyses of averaged DNAm changes are concordant with prior reports of both averaged DNAm and gene expression.

For this study, we took the novel approach of whole genome analysis followed by a targeted strategy using a set of candidate genes developed from prior reports of n-3 PUFAs, inflammation, and breast cancer effects to test for inflammation-related DNAm changes. The RRBS experiment uses bisulfite conversion which is the gold standard for DNAm assays. Our focused anlaysis on a hypothesis-derived set of candidate genes was conducted with a genome-wide statistical cutoff which minimizes false positives while still focusing on inflammation. As the first opportunity to test the capability of our candidate genes to detect n-3 PUFA treatment effects, this study showed that the candidate genes were enriched in DNAm differences after n-3 PUFA treatment. To the best of our knowledge, the six-month n-3 PUFA treatment of 5 g/day EPA+DHA resulted in detection of more gene associated DMCs than any other reported n-3 PUFA intervention to date. In order to understand how the large number of observed n-3 PUFA specific DNAm changes involved inflammation signaling, we focused on pathways that involved our candidate genes and featured directionally concordant DNAm changes.[32] This approach identified at least two hypermethylated pathways involved in regulating inflammation with biologically relevant localization of DMCs with hypermethylation observed after n-3 PUFA treatment consistent with decreased inflammation or inflammatory potential: Focal Adhesion and TLR pathways. The TLR pathway is interesting both for effects on immune response and production of pro-inflammatory cytokines; additionally, prior reports have demonstrated modulation of TLR signaling by n-3 PUFAs.[4, 18, 25, 37] Likewise, previous research has shown n-3 PUFA modulation of gene expression in the focal adhesion pathway[8] and of DNAm in the PI3K signaling component of the focal adhesion pathway.[47]. Such concordance lends support to the findings of our study and analytic approach.

The limitations of this study relate primarily to the small sample size. Although 10 paired samples are sufficient for detection of average DNAm changes due to dietary n-3 PUFAs, a larger study population is needed to explore the variability of DNAm changes with n-3 PUFA treatment as well as the interplay between DNAm and biological parameters. Despite the potential inaccuracy of estimating cell composition of PBMC specimens via indirect method of Hicks et al, we used the estimated cell populations to control for the possibility that the observed DNAm changes were caused by cell type changes. However, analysis of different cell populations for each patient specimen was outside the purview of this study. Analysis of DNAm of PBMCs from another dose group, such as 0.84 g/d n-3 PUFAs, would have also provided a useful comparison group; however, our previous dose finding study could not statistically differentiate between 3-7 g per day arms.[53] Therefore, these results will inform future n-3 PUFA studies with interventions that achieve similar DHA, EPA levels1 Additionally, we acknowledge the potential limitations of focusing on a predetermined set of candidate genes, combined with directional enrichment of DNAm in selection of pathways; however, this analytic approach enabled identification of two inflammation-related signaling networks that have also been reported by others as responsive to n-3 PUFAs. Finally, changes in DNAm may not indicate functional regulation of gene expression even when occurring in the promoter region.[16] We chose a broad definition of the promoter region and looked for concerted DNAm changes in pathways to increase the probability of identifying pathways under epigenetic regulation. Future studies will address the functional relationship between DNAm and gene expression and biologic response of n-3 PUFA treatment.

Taken together, our study results demonstrate the impact of a six-month, high dose regimen of n-3 PUFAs in women at high risk of breast cancer by assaying DNAm genome-wide in PBMCs. While changes in average global DNAm were not apparent, our analyses of DMCs indicated hypermethylation of key pro-inflammatory signal transduction pathways that include PI3K and TLR signaling. Further research is warranted to evaluate the potential for individual responses in PBMC DNAm and the implications for monitoring and tailoring n-3 PUFA intake.

## Data Availability

Processed data is available upon request

## Acknowledgments

LDY, DEF, PSY, and RAB designed the research; MS, SW, and DEF conducted research; DEF analyzed data and performed statistical analysis; DEF and LDY wrote the paper; LDY had primary responsibility for final content. All authors read and approved the final manuscript.

