## Supplemental Material for "Peripheral Blood DNA Methylation Changes after Omega-3 Fatty Acid Treatment Indicate Anti-inflammatory Effects and Individual Variability"

| | Undosed Average | Dosed Average | Paired T-test Pvalue |
|---|---|---|---|
| *Fatty Acid Summary* | | | |
| **Saturated FA** | **32.79%** | **33.29%** | **0.54** |
| **Monounsaturated FA** | **22.12%** | **16.70%** | ***0.0011*** |
| **n-3 PUFA** | **4.59%** | **14.49%** | ***0.0000067*** |
| **n-6 PUFA** | **39.78%** | **34.72%** | ***0.011*** |
| **n3:n6 Ratio** | **0.12** | **0.42** | ***0.0000086*** |
| *Individual Fatty Acids* | | | |
| C14:0 | 0.76% | 0.66% | 0.51 |
| C16:0 | 22.39% | 22.13% | 0.68 |
| C16:1n7 | 2.46% | 1.07% | 0.25 |
| C16:3n4 | 0.17% | 0.10% | *0.058* |
| C18:0 | 9.51% | 10.46% | *0.077* |
| C18:1n9 | 17.56% | 13.87% | *0.00030* |
| C18:1n7 | 1.96% | 1.66% | *0.052* |
| C18:2n6 | 28.51% | 26.17% | 0.22 |
| C18:3n6 | 0.45% | 0.27% | *0.000092* |
| C18:3n3 | 0.55% | 0.49% | 0.60 |
| C9-T11 CLA | 0.55% | 0.71% | 0.25 |
| C20:0 | 0.14% | 0.04% | 0.47 |
| C20:1n9 | 0.13% | 0.10% | 0.16 |
| C20:2n6 | 0.27% | 0.27% | 1.00 |
| C20:3n6 | 2.12% | 1.28% | *0.0028* |
| C20:4n6 | 8.12% | 6.63% | *0.0047* |
| C20:4n3 | 0.23% | 0.19% | 0.68 |
| C20:5n3 EPA | 0.92% | 6.38% | *0.0000058* |
| C22:2n6 | 0.03% | 0.03% | 0.99 |
| C22:4n6 | 0.28% | 0.08% | *0.00069* |
| C22:5n3 | 0.72% | 1.32% | *0.0018* |
| C22:6n3 DHA | 2.17% | 6.11% | *0.00030* |

**Supplemental Table 1: Change in Serum Fatty Acids**

Fatty acid content is averaged before and after n-3 PUFA treatment as both type of fatty acid (top portion of the chart) and individual fatty acid (bottom portion of the chart). Changes in fatty acids were determined using a paired T-test which is reported in the last column. Fatty acids are reported as a percent of total fatty acids.

| Genes | Reference |
|---|---|
| ADORA3, AHSG, ALOX15, ALOX5, ALOX5AP, ANXA1, AOAH, AOX1, ATRNL1, BCL6, BDKRB1, C3, C3AR1, CCL1, CCL11, CCL13, CCL2, CCL23, CCL24, CCL26, CCL5, CCL7, CCL8, CCR1, CCR2, CCR3, CCR7, CD40, CD74, CRP, CXCL1, CXCL12, CXCL2, CXCL3, CXCL5, CXCL6, DOCK2, EPHX2, F11R, FPR1, GPR68, HDAC4, HDAC9, HRH1, IL17C, IL18RAP, IL1A, IL1B, IL1R1, IL20, IL31RA, IL8RB, IRAK2, IRF7, KLKB1, KNG1, LTB4R, LY75, LY86, LY96, NFAM1, NFATC3, NFATC4, NFE2L1, NMI, NOS2A, NR3C1, PARP4, PF4, PLA2G7, PREX1, PRG2, PROC, PROK2, PTGS2, PTX3, S100A12, S100A8, S100A9, SARM1, SCUBE1, SELE, TICAM2, TLR1, TLR4, TLR5, TNFAIP6, TOLLIP, TPST1 | GDAC 2012 Analysis[1] |
| Cox2, VEGF, ERK1/2, NF-Kb | De Caterina 2005[2] |
| CPB2, DUSP2, EBI3, SIAHBP1 | Hammamieh 2007[3] |
| EZH2, IGFBP3, CDH1 | Dimri 2009[4] |
| HMGCS2 | De Rosa 2015[5] |
| SCD, FADS2 | Gillies 2012[6] |
| HIF1A, CREB1 | Tsunoda 2015[7] |
| TNFa, DNMT1, DNMT3B, IL6 | Cormier 2014[8], Li 2012[9] |
| CD36, FFAR3, CD14, PDK4, FADS1 | Iira do Amaral 2014[10] |
| CRMP1, GDNF, GFRA1, MYL9, ROBO1, ROBO3, SEMA5A, C9orf125, COL14A1, ENPP2, ERG2, PLD5, ROBO3, RUNX1T1, SEMA5A, TBX18, TSHZ3, ZBTB16, ZNF208, SLC6A3, C6orf174, ZNF254, DMRTA2, LHX8, WT1, WT1-AS, HOXB13, ECEL1, SOX2-OT | Stirzaker 2015[11] |
| RASSF1 | Yan 2006[12] |
| RASSF1-AS, RARB | Antil 2010[13] |
| CDKN2A | Bean 2007[14] |
| EGFR | Schley 2007[15] |
| GPX3 | Chen 2011[16], Mohamed 2014[17] |
| EIB3, FOX3P | Hammamieh 2007[3] |
| DNMT1, CXCL2, IL-1β, NOS2, TNF-α, DNMT3a | Niwa 2012[18], Nakano 2013[19] |

**Supplemental Table 2: Candidate Genes**

Selection of the 152 candidate genes was based on either: 1) inflammation related genes from GDAC analysis of the TCGA breast cancer dataset or 2) genes involved in fatty acid metabolism, inflammation, and/or breast cancer.

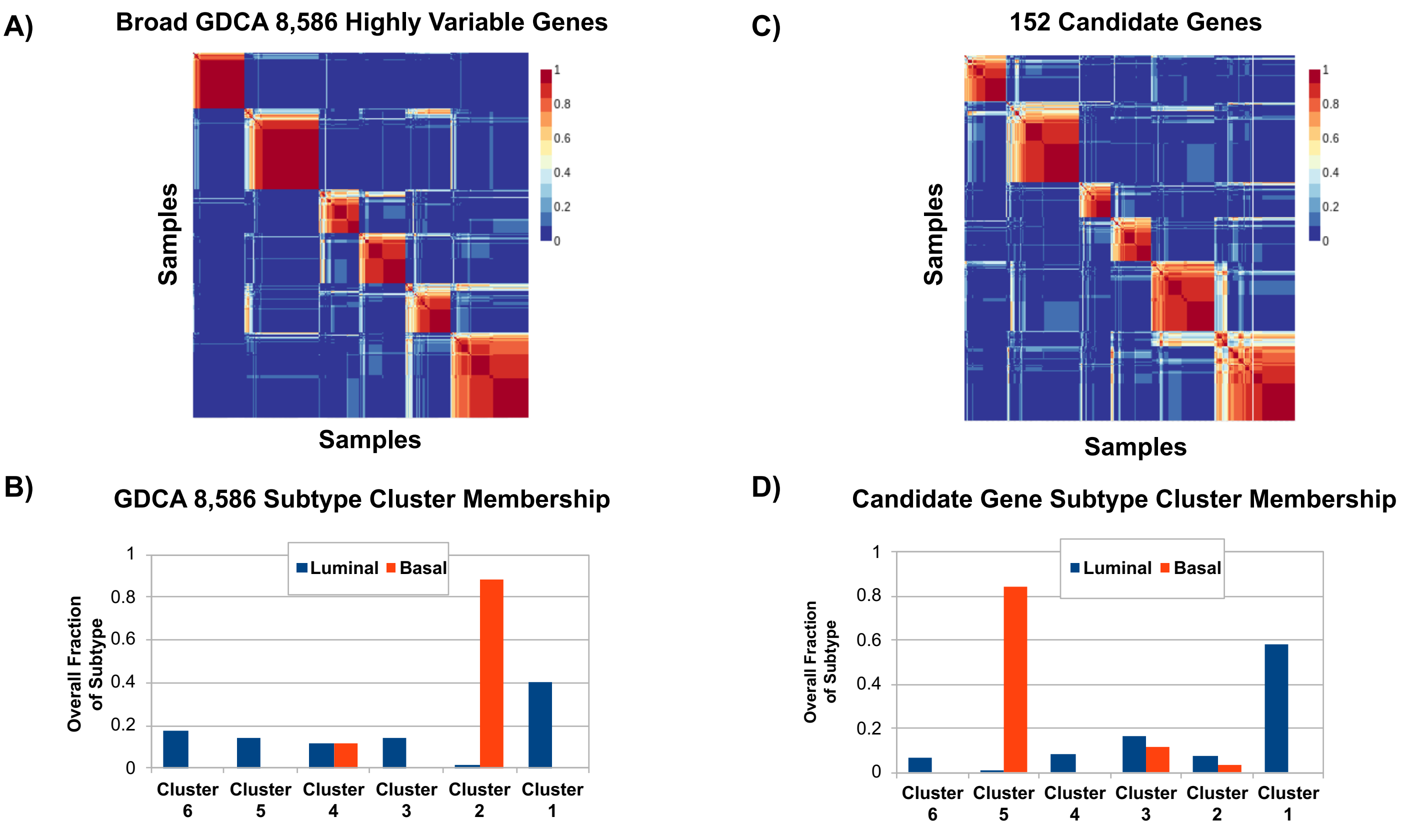


**Supplemental Figure 1: DNAm of Candidate Genes Stratifies TCGA Breast Cancer Samples similar to GDAC Analysis**

A) Non-negative matrix factorization (NMF) clustering was produced using 6 groups on the GDAC derived 8,586 most variable genes among the TCGA breast cancer cohort. B) Comparison of samples in each cluster shows that luminal and basal subtypes dominate two of the clusters identified using the 8,586 genes identified by the GDAC analysis. The y-axis shows the fraction of all TCGA samples represented in each cluster (Note: only samples identified as Luminal or Basal are reported in B) whereas A) shows all samples used in GDAC analysis). C) NMF clustering using 6 groups on the 152 candidate genes performed on the TCGA breast cancer cohort. Clustering using only the 152 candidate genes produced a similar sample grouping when compared to the GDAC analysis with 8,586 genes. D) Candidate gene clusters show that luminal and basal subtypes also dominate two clusters.

| Gene | Chromosome | CpG Position | P-value | Q-value | DNAm Difference (%) |
|---|---|---|---|---|---|
| **ALOX15** | chr17 | 4639431 | 7.0E-04 | 1.6E-02 | -8 |
| **ALOX5** | chr10 | 45374653 | 2.0E-03 | 3.2E-02 | -5 |
| **ALOX5** | chr10 | 45374654 | 6.8E-04 | 1.6E-02 | 5 |
| **AOX1** | chr2 | 200586226 | 9.6E-04 | 2.0E-02 | -7 |
| **CDKN2A** | chr9 | 21994543 | 4.0E-08 | 5.3E-06 | 6 |
| **CREB1** | chr2 | 207529992 | 2.6E-04 | 7.9E-03 | 10 |
| **CRMP1** | chr4 | 5890886 | 1.5E-03 | 2.6E-02 | 6 |
| **CXCL12** | chr10 | 44385636 | 2.8E-04 | 8.5E-03 | -9 |
| **DMRTA2** | chr1 | 50419460 | 9.4E-06 | 6.1E-04 | 10 |
| **ECEL1** | chr2 | 232490585 | 3.3E-04 | 9.4E-03 | 8 |
| **EZH2** | chr7 | 148885245 | 6.5E-08 | 8.3E-06 | 8 |
| **EZH2** | chr7 | 148885258 | 7.6E-04 | 1.7E-02 | 6 |
| **FADS2** | chr11 | 61827576 | 2.9E-04 | 8.6E-03 | 6 |
| **FASN** | chr17 | 82103451 | 1.2E-07 | 1.4E-05 | 7 |
| **FASN** | chr17 | 82097466 | 3.8E-03 | 4.8E-02 | -5 |
| **FFAR3** | chr19 | 35352196 | 4.1E-09 | 6.9E-07 | -17 |
| **FFAR3** | chr19 | 35352213 | 6.5E-09 | 1.1E-06 | -17 |
| **FFAR3** | chr19 | 35352218 | 1.7E-08 | 2.5E-06 | -16 |
| **FFAR3** | chr19 | 35352204 | 8.1E-07 | 7.7E-05 | -14 |
| **HOXB13** | chr17 | 48734626 | 2.9E-03 | 4.0E-02 | 9 |
| **IGFBP3** | chr7 | 45921153 | 2.8E-04 | 8.3E-03 | 6 |
| **IL17C** | chr16 | 88637610 | 1.2E-03 | 2.2E-02 | 9 |
| **IRF7** | chr11 | 615109 | 7.1E-04 | 1.6E-02 | -6 |
| **LEP** | chr7 | 128241029 | 1.8E-04 | 6.0E-03 | -7 |
| **LHX8** | chr1 | 75130134 | 8.4E-04 | 1.8E-02 | -11 |
| **LHX8** | chr1 | 75125505 | 6.8E-04 | 1.6E-02 | 7 |
| **LHX8** | chr1 | 75125474 | 3.9E-04 | 1.0E-02 | 6 |
| **MYL9** | chr20 | 36541768 | 2.9E-13 | 1.0E-10 | -18 |
| **NOS2** | chr17 | 27808814 | 1.3E-04 | 4.9E-03 | 11 |
| **PREX1** | chr20 | 48833850 | 6.7E-09 | 1.1E-06 | 11 |
| **RARB** | chr3 | 24829608 | 1.1E-03 | 2.1E-02 | -6 |
| **RASSF1-AS1** | chr3 | 50328541 | 3.9E-03 | 4.8E-02 | 13 |
| **RUNX1T1** | chr8 | 92102072 | 2.1E-04 | 6.8E-03 | 8 |
| **RUNX1T1** | chr8 | 92103027 | 2.8E-06 | 2.2E-04 | 8 |
| **RUNX1T1** | chr8 | 92103035 | 2.7E-05 | 1.4E-03 | 6 |
| **S100A8** | chr1 | 153394933 | 2.5E-05 | 1.3E-03 | 6 |
| **SLC6A3** | chr5 | 1446597 | 2.0E-03 | 3.2E-02 | 5 |
| **TBX18** | chr6 | 84773926 | 1.3E-06 | 1.1E-04 | -5 |
| **TOLLIP** | chr11 | 1310191 | 8.4E-04 | 1.8E-02 | 12 |
| **TOLLIP** | chr11 | 1310213 | 3.0E-04 | 8.7E-03 | 12 |
| **TOLLIP** | chr11 | 1310233 | 2.6E-03 | 3.8E-02 | 10 |
| **TOLLIP** | chr11 | 1306787 | 4.6E-06 | 3.4E-04 | 10 |
| **TPST1** | chr7 | 66205409 | 4.7E-05 | 2.2E-03 | -6 |
| **VEGFA** | chr6 | 43771228 | 2.5E-03 | 3.7E-02 | 6 |
| **WT1** | chr11 | 32430231 | 2.2E-03 | 3.3E-02 | 12 |
| **WT1** | chr11 | 32435765 | 3.9E-03 | 4.8E-02 | 9 |
| **WT1-AS** | chr11 | 32430231 | 2.2E-03 | 3.3E-02 | 12 |
| **WT1-AS** | chr11 | 32435765 | 3.9E-03 | 4.8E-02 | 9 |
| **ZNF208** | chr19 | 22010923 | 1.5E-03 | 2.6E-02 | -6 |

**Supplemental Table 3: Candidate Gene Promoter DMCs**

Differentially methylated CpGs (DMCs) of candidate gene promoters were identified using MethylKit v1.2.0 which tested for DNAm change between the untreated and treated samples for all pass filter CpGs (≥10 reads coverage in ≥ 16 samples). A CpG was considered a DMC if the multiple test corrected P-value (Q-value) was less than 0.05 and the average DNAm difference between the untreated and the treated samples was at least 5%. Here, the candidate gene promoter DMCs were identified by intersecting the candidate genes with all promoter DMCs.

| Term | Overlap | P-value | Adjusted P-value | Genes |
|---|---|---|---|---|
| Photodynamic therapy-induced HIF-1 survival signaling | 3/37 | 0.00003 | 0.00379 | NOS2; IGFBP3; VEGFA |
| Effects of Nitric Oxide | 2/8 | 0.00007 | 0.00469 | NOS2; AOX1 |
| Vitamin D Receptor Pathway | 4/182 | 0.00020 | 0.00886 | CDKN2A; ALOX5; IGFBP3; S100A8 |
| Circadian rhythm related genes | 4/201 | 0.00029 | 0.00971 | CREB1; NOS2; LEP; EZH2 |
| Transcription factor regulation in adipogenesis | 2/22 | 0.00056 | 0.01415 | CREB1; LEP |
| Senescence and Autophagy in Cancer | 3/105 | 0.00062 | 0.01415 | CDKN2A; IGFBP3; IRF7 |
| Tumor suppressor activity of SMARCB1 | 2/31 | 0.00112 | 0.01905 | CDKN2A; EZH2 |
| Toll-like Receptor Signaling | 2/31 | 0.00112 | 0.01905 | TOLLIP; IRF7 |
| Bladder Cancer | 2/40 | 0.00186 | 0.02814 | CDKN2A; VEGFA |
| Structural Pathway of Interleukin 1 (IL-1) | 2/49 | 0.00278 | 0.03154 | TOLLIP; IRF7 |
| Hepatitis C and Hepatocellular Carcinoma | 2/49 | 0.00278 | 0.03154 | NOS2; VEGFA |
| NO/cGMP/PKG mediated Neuroprotection | 2/47 | 0.00256 | 0.03154 | CREB1; NOS2 |
| G1 to S cell cycle control | 2/64 | 0.00470 | 0.03895 | CREB1; CDKN2A |
| Oncostatin M Signaling Pathway | 2/65 | 0.00484 | 0.03895 | CREB1; VEGFA |
| MECP2 and Associated Rett Syndrome | 2/62 | 0.00442 | 0.03895 | CREB1; EZH2 |
| RIG-I-like Receptor Signaling | 2/60 | 0.00414 | 0.03895 | CXCL12; IRF7 |
| Non-small cell lung cancer | 2/66 | 0.00499 | 0.03895 | CDKN2A; RARB |
| Sterol Regulatory Element-Binding Proteins (SREBP) signaling | 2/69 | 0.00544 | 0.03895 | CREB1; FASN |
| AMP-activated Protein Kinase (AMPK) Signaling | 2/69 | 0.00544 | 0.03895 | LEP; FASN |
| Leptin signaling pathway | 2/76 | 0.00657 | 0.04465 | CREB1; LEP |
| SCFA and skeletal muscle substrate metabolism | 1/6 | 0.00956 | 0.04645 | FFAR3 |
| Robo4 and VEGF Signaling Pathways Crosstalk | 1/6 | 0.00956 | 0.04645 | VEGFA |
| Nicotine Metabolism | 1/6 | 0.00956 | 0.04645 | AOX1 |
| Pancreatic adenocarcinoma pathway | 2/89 | 0.00891 | 0.04645 | CDKN2A; VEGFA |
| Allograft Rejection | 2/89 | 0.00891 | 0.04645 | CXCL12; VEGFA |
| Pathways in clear cell renal cell carcinoma | 2/85 | 0.00815 | 0.04645 | FASN; VEGFA |
| Apoptosis Modulation and Signaling | 2/91 | 0.00929 | 0.04645 | CDKN2A; TOLLIP |
| Apoptosis | 2/84 | 0.00797 | 0.04645 | CDKN2A; IRF7 |
| MicroRNA for Targeting Cancer Growth and Vascularization in Glioblastoma | 1/7 | 0.01115 | 0.04891 | VEGFA |
| EV release from cardiac cells and their functional effects | 1/7 | 0.01115 | 0.04891 | CXCL12 |
| Molybdenum cofactor (Moco) biosynthesis | 1/7 | 0.01115 | 0.04891 | AOX1 |

**Supplemental Table 4: Significantly Enriched Pathways using Candidate Gene Promoter DMCs**

Candidate genes that contained a DMC in their promoter region were used to determine which pathways were significantly enriched for DNAm changes after n-3 PUFA treatment. Overlap provides the number of candidate gene promoter DMCs that were found in the pathway out of the total number of genes in the pathway.

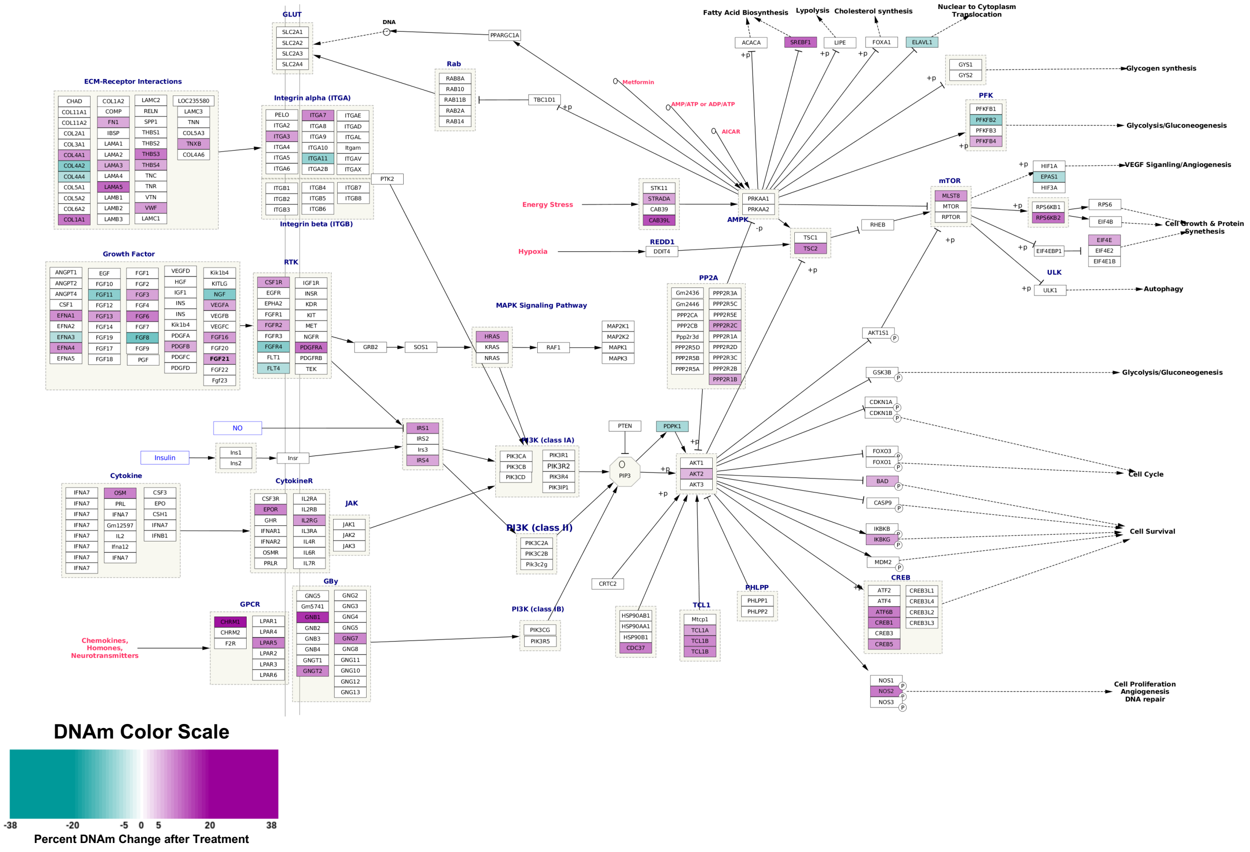


***Supplemental Figure 2:*** **Focal Adhesion-PI3K-Akt-mTOR-signaling pathway**

A) The Focal Adhesion-PI3K-Akt-mTOR-signaling pathway was found to be significantly enriched for both candidate gene promoter DMCs and for hypermethylation of genome-wide promoter DMCs (hypergeometric FDR p-value = 0.023). Only CpGs where the magnitude of the individual's DNAm change was greater than 5% were counted so that the variability was not overrepresented by small changes. Hypermethylated DMCs are localized to the cytokine and chemokine receptors and the AKT portion of the pathway. Pathway was constructed using WikiPathways and Cytoscape.

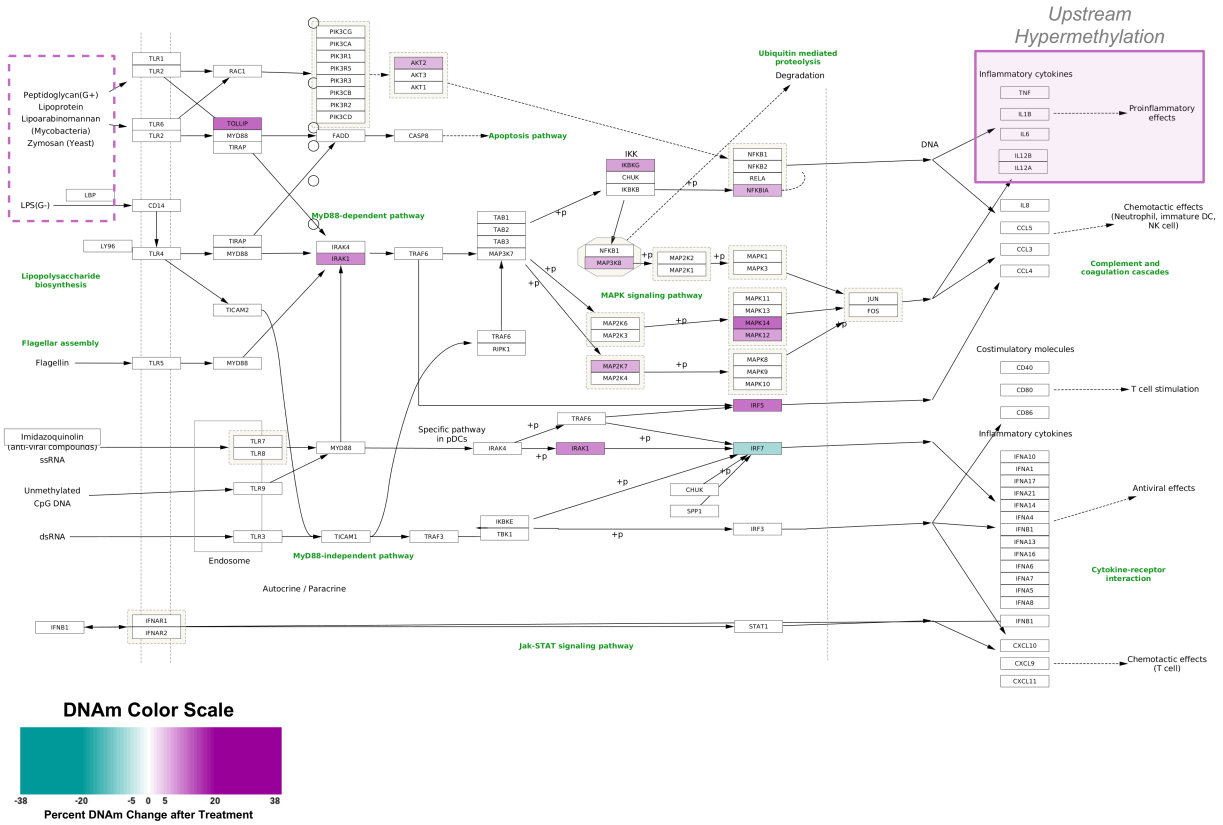


### Supplemental Figure 3: Toll-Like Receptor Signaling Pathway

A) The Toll-like Receptor Signaling Pathway was found to be significantly enriched for both candidate gene promoter DMCs and for hypermethylation of genome-wide promoter DMCs (hypergeometric FDR p-value = 0.029). Only CpGs where the magnitude of the individual's DNAm change was greater than 5% were counted so that the variability was not overrepresented by small changes. Hypermethylation occurs downstream of TLR1, 2, 4, 5, and 6 and upstream of the pro-inflammatory cytokines. Pathway was constructed using WikiPathways and Cytoscape.

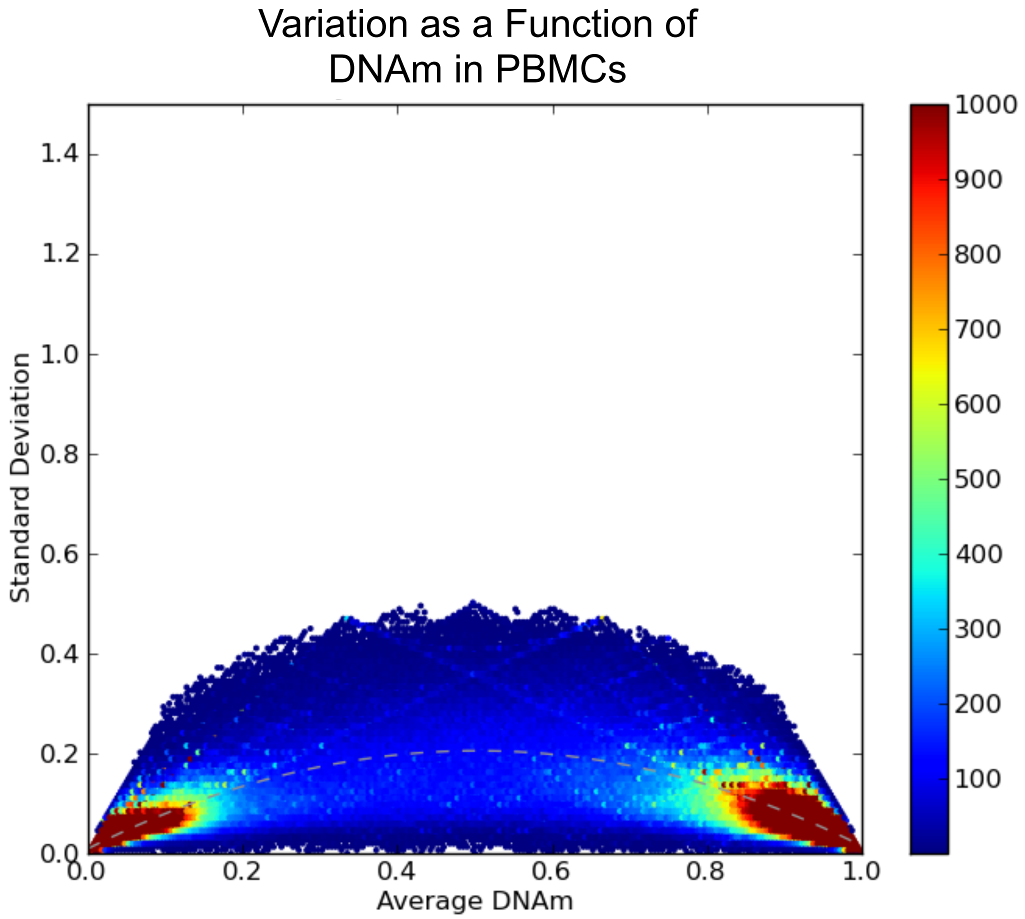


**Supplemental Figure 4: Variation as a Function of DNAm in PBMCs**

The variation in DNAm between samples was calculated in order to determine the power of detecting DNAm changes. The variation of DNAm for all participants was plotted as function of DNAm and fit to a parabola (grey dashed line). The variation of DNAm in our PBMC samples as assayed by RRBS was lower (max of fit SD = 20%) than the variation determined using publicly available human PBMC data[20] (GSE57107) that was assayed using the Infinium 450k Array (max of fit = 31%; data not shown).

**REFERENECES**
